# Comparison of Shoulder Functions Between Workers With and Without Shoulder Impingement Syndrome

**DOI:** 10.1101/2020.05.24.20109736

**Authors:** Jun-hee Kim, Oh-yun Kwon, Chung-hwi Yi, Hye-seon Jeon, Woo-chol Joseph Choi, Jong-hyuck Weon

## Abstract

The occurrence of shoulder impingement syndrome (SIS) is associated with the frequent handling and lifting of heavy loads and excessive repetitive work above the shoulder level. Thus, assembly workers have a high prevalence of shoulder injuries, including SIS. The purpose of this study was to investigate differences in shoulder ROM, muscle strength, asymmetry ratio, function, productivity, and depression between workers with and without SIS.

Sixty-seven assembly line male workers (35 workers with SIS and 32 workers without SIS) participated in this study. The four shoulder ROMs and the five muscle strengths were measured using a Smart KEMA system. The asymmetry ratios were calculated using the asymmetry ratio formula; shoulder functions were measured using the shoulder pain and disability index (SPADI), disabilities of the arm, shoulder, and hand (DASH), and visual analogue scale (VAS); and Endicott work productivity scale (EWPS). Severity of depression was measured using the Beck depression inventory (BDI). Independent t-tests were performed for statistical analysis.

The SPADI, DASH, and VAS values of workers with SIS were significantly higher than those of workers without SIS. Also, workers with SIS had significantly smaller shoulder internal rotation and shoulder abduction compared to workers without SIS. In addition, workers with SIS exhibited significantly lower SIR muscle strength than workers without SIS. Workers with SIS had significantly higher asymmetry ratios in shoulder internal rotation, shoulder external rotation, and elbow flexion muscle strength than workers without SIS.

The SPADI and DASH scores, which indicate shoulder function, were higher, and the intensity of self-aware pain was higher in workers with SIS. Also, workers with SIS exhibited reduced SIR and SAB ROMs; decreased SIR muscle strength. Particularly, the asymmetry ratios of SIR, SER, EF muscle strength are good comparable factors for workers with and without SIS. In addition, the asymmetry ratios of shoulder muscle strengths could provide an important baseline comparison for the workers with SIS.

## Introduction

Shoulder impingement syndrome (SIS) is common not only among athletes performing overhead movements such as throwing or swimming but also among workers who perform constant arm movements [1,2]. Especially, intensive work using shoulder joints can be a risk factor for SIS in workers [1]. Repeated strain on the shoulder joint and working with hands above the acromion height can lead to mechanical compression of the rotator cuff, subacromial bursa, and biceps tendon, leading to shoulder disease such as tendinitis, bursitis, and rotator cuff tear [1].

SIS considered to be a part of the process involved in the degeneration of the rotator cuff [3,4]. Therefore, early identification of the physical factors that could mediate SIS is important [3]. Several factors contribute to the onset of SIS, including abnormal acromial morphology, incorrect kinematic patterns associated with altered rotator cuff or scapular muscle function, and capsular abnormalities (including posterior capsular tightness and capsular laxity) [2,5,6].

Studies have suggested that SIS patients exhibit decreased range of motion (ROM) of the shoulder joint [2,7]. Warner et al. (1990) found that patients with SIS exhibited significantly reduced shoulder internal rotation (SIR) and shoulder horizontal adduction (SHA) angle compared to those with shoulder instability. Similarly, Tyler et al. (2000) found that patients with SIS in the dominant arm exhibited reduced SIR ROM compared to normal subjects, and patients with SIS in the non-dominant arm exhibited reduced SIR and shoulder external rotation (SER) ROM. In addition, patients with SIS showed increased posterior capsule tightness. Therefore, Tyler et al. (2000) suggested that a decrease in the SIR ROM of patients with SIS is associated with posterior capsule tightness.

The shoulder complex, composed of the clavicle, scapula, and humerus, relies on muscles to provide dynamic stability during its wide ROM [8]. Proper balance among the muscles surrounding the shoulder complex is essential for shoulder mobility [9,10]. The lack of ROM or strength in specific muscles must be compensated for by another shoulder muscle, which leads to shoulder dysfunction [11]. These muscle imbalances lead to changes in joint kinematics and, eventually, SIS [11]. Dr. Janda suggested that weakness of shoulder muscles, such as the serratus anterior, infraspinatus, lower and middle trapezius, as well as tightness of the pectorals and upper trapezius muscles, cause SIS [9]. Other studies have reported that weakness of the rotator cuff muscles affects the onset of SIS [12,13].

Although the relative weakness or tightness of certain muscles among these scapulo-thoracic and scapulo-humeral muscles may cause SIS, other studies have reported conflicting results. According to Erol et al. (2008), the strength of SIR did not differ significantly between people with and without SIS. Likewise, no significant difference was found in terms of in SER strength between the people with and without SIS [14]. Only a deficit in relative SIR muscle strength on the involved side compared to that on the normal side was found to be a significant difference between those with and without SIS [14]. Bak and Magnusson (1997) reported no significant difference in the SER strength of elite swimmers with and without SIS.

Although many studies have compared physical factors such as shoulder ROM and muscle strength in patients with and without SIS, no study has compared the physical, functional, and psychological factors of workers with and without SIS. Therefore, this study aimed to identify any differences in shoulder ROM, muscle strength, asymmetry ratio, function, pain, productivity, and severity of depression between workers with and without SIS.

## Materials and Methods

### 1. Subjects

Sixty-seven assembly line male workers—35 workers with SIS and 32 workers without SIS—participated in this study. All subjects were explained the risks and benefits associated with this study, and they signed an informed consent form. Workers with SIS were included if they had a positive sign in two or more SIS screening tests and in two of the specific SIS tests. SIS screening and specific SIS tests were conducted by a physical therapist with 5 years of experience in physical therapy. The SIS screening tests were as follows: (a) persistent pain in the proximal anterior or lateral shoulder for more than 1 week within the previous 6 months; (b) a painful arc during active shoulder elevation; (c) tenderness upon palpation of the rotator cuff tendons; and (d) pain during resisted isometric shoulder abduction (SAB) at 90° or a positive Jobe’s test (empty can test) [16]. The specific SIS tests used in this study were the Neer and Hawkins tests [17]. The exclusion criteria were history of dislocation or traumatic injuries of the shoulder joint or history of shoulder joint surgery. This study was approved by the Yonsei University Mirae institutional review board (1041849-201710-BM-112-02).

### 2. Experimental Instruments

#### 2.1 Smart KEMA Motion Sensor

A Smart KEMA motion sensor (Smart KEMA motion sensor, KOREATECH Co., Ltd., Seoul, Korea) was used to measure the kinematics of SIR, SER, SAB, and SHA. The Smart KEMA motion sensor outputs all shoulder ROM measurements in degrees. The motion sensor was mounted on a plastic frame and placed in a strap to fix them in place. This motion sensor consists of a tri-axillar gyroscope, magnetometer, and an accelerometer, as well as a signal converter and signal transmission sensor. Motion sensor data were transmitted to a recording Android tablet with Smart KEMA software, at a sampling frequency of 25 Hz. The value of the real-time angle for 5 s was recorded at the end of ROM, and the Smart KEMA software computed the average of the angle data of in the middle 3 s for data analysis.

#### 2.2 Smart KEMA Tension Sensor

The isometric muscle strength of SIR, SER, elbow flexion (EF), lower trapezius (LT), and scapular protraction (SP) were measured using a Smart KEMA tension sensor (Smart KEMA tension sensor, KOREATECH Co., Ltd., Seoul, Korea). The tension sensor contained a load cell with a measurement range of 0–1,960 N and an accuracy of ± 4.9 N. The tension sensor data were transmitted at a sampling frequency of 10 Hz to a recording Android tablet running Smart KEMA software. The real-time value of strength for was recorded over 5 s, and the Smart KEMA software computed the average of the data for the middle 3 s for data analysis. In previous studies, this tension sensor was used to measure isometric strength, and it exhibited high intra-rater reliability [18,19]. The tension sensor had two rings. One side of the sensor was fixed to a glass suction cup or a stable material by using an orthopedic belt and the other side was fixed to a body segment by using the strap.

### 3. Outcome Measures

#### 3.1 Visual Analogue Scale

The visual analogue scale (VAS) was used to measure the severity and perception of shoulder pain caused by SIS. VAS ranges from 0 mm (no pain) to 100 mm (worst pain imaginable). The subjects were asked to record their greatest pain during daily activities conducted using the affected arm for 1 week.

#### 3.2 Shoulder Pain and Disability Index

The shoulder pain and disability index (SPADI) that was revised for Koreans was used to evaluate the symptoms and severity of SIS [20]. The questionnaire contained 13 self-reporting items, of which five items belonged to the pain subscale and eight to the function and disability subscale. The total score of this tool is determined by averaging the scores of the 13 evaluation items. The severity of shoulder pain and disability increase as the average score increases from 0 to 100, with zero being the least painful and 100 the most painful.

#### 3.3 Disabilities of the Arm, Shoulder, and Hand

The disabilities of the arm, shoulder, and hand (DASH) that was revised for Koreans was used to evaluate the subjective state of the upper limbs [21]. The DASH disability and symptom score is determined based on 30 items. The higher the score, the more severe is the impairment [22].

#### 3.4 Endicott Work Productivity Scale

The self-administered Endicott work productivity scale (EWPS) questionnaire consists of 25 items, and it assesses the extent to which an individual’s medical condition affects their work performance [23].

#### 3.5 Beck Depression Inventory

The Beck depression inventory (BDI) is a 21-question multiple-choice self-report inventory, and it is among the most widely used psychometric tests for measuring depression severity. In this study, the Korean-BDI version I was used [24]. Numerical values of 1-4 are assigned to each statement to indicate the degree of depression severity [25].

#### 3.6 Measurement of Shoulder ROM

##### 3.6.1 Shoulder internal rotation

To measure SIR, subjects were positioned supine on a table, with the shoulder to be tested positioned at 90° of glenohumeral abduction. Manual scapular stabilization was achieved by applying a posteriorly directed force against the subject’s coracoid process and clavicle with the heel of the hand to prevent anterior translation of the subject’s humeral head [26,27].

##### 3.6.2 Shoulder external rotation

To measure SER, subjects were positioned supine on a table with the shoulder to be tested positioned at 90° of glenohumeral abduction. Manual scapular stabilization was achieved by applying a posteriorly directed force against the subject’s coracoid process and clavicle with the heel of the hand to prevent anterior translation of the subject’s humeral head [28].

##### 3.6.3 Shoulder abduction

SAB ROM measurement was performed in a sitting position. In the sitting position on the table, the subject’s palm was pointed along the forward direction. To avoid SHA in the forward direction when performing SAB, the examiner held the subject’s distal humerus and performed the abduction motion until the end feel of the shoulder joint was felt.

##### 3.6.4 Shoulder horizontal adduction

To measure SHA, the shoulder to be measured was placed on the table with the subject lying down sideways to prevent abduction of the subject’s scapula during the movement. The frontal plane of the body was aligned to be perpendicular to the ground. Then, the shoulder was flexed by 90°, such that the subject’s palm faced the ceiling. The examiner held the distal part of the humerus and moved it in the horizontal adduction direction without EF, while the upper body or acromion was prevented from rotating in the front or rear direction [29]. SHA ROM was measured when the examiner felt the end feel of the shoulder joint.

#### 3.7 Measurement of Shoulder Muscle Strength

##### 3.7.1 Shoulder internal rotation

To measure the isometric strength of SIR, subjects were asked to maintain the shoulder in the neutral position and flex the elbow at 90° in a side-lying position with the test arm facing the table. Then, the wrist of the subject was moved toward the ceiling, and SIR was performed with the maximum force. To prevent compensatory motion of trunk rotation by the subject, the examiner fixed the trunk of the subject by hand.

##### 3.7.2 Shoulder external rotation

To measure the isometric strength of SER, subjects flexed their shoulder and elbow at 90° in a side-lying position with the test arm facing the ceiling. Then, the wrist of the subject was moved toward the ceiling, and SER was performed with the maximum force. To prevent the compensatory motion of shoulder horizontal abduction, the subject was instructed to not move the tested upper arm away from the palm of the opposite hand [30].

##### 3.7.3 Elbow flexion

To measure the isometric strength of EF, each subject was asked to sit in an upright position at the edge of a table. The subjects performed EF against a strap. To prevent shoulder flexion and elevation during EF, the subjects held their distal part of upper arm by hand to fix the upper arm along the side of the trunk.

##### 3.7.4 Lower trapezius

The isometric strength of the LT muscle was measured by the subject in a prone position. The subject abducted the shoulder at 120°, fully extended the elbow joints, and rotated the shoulder externally as far as possible [31]. The subject performed the motion by applying the maximum force with the wrist directed toward the ceiling.

##### 3.7.5 Scapular protraction

The isometric strength of SP was measured in the supine position. The subject flexed the shoulder joint at 90°. The subject performed SP with the maximum force toward the ceiling. During strength measurement, the subject’s trunk was fixed with the opposite hand to ensure that the subject could not rotate their trunk.

#### 3.8 Asymmetry ratio of shoulder muscle strength

The asymmetry ratio between the uninvolved and involved sides was calculated using the following formula: percent asymmetry ratio (%AR) of shoulder muscle strength = (uninvolved − involved side)/(uninvolved + involved side) * 100 [30]. To calculate the percent asymmetry ratio of shoulder muscle strength, the uninvolved side was replaced with the dominant side in the asymmetry ratio formula when comparing the normal worker group to the SIS group with an uninvolved side, and the non-dominant side was replaced with the involved side when comparing the normal worker group to the SIS group with an involved side [7,30,32].

### 4. Experimental Procedures

Before data collection, the subjects were familiarized with the testing protocol, provided instructions, and asked to practice the shoulder ROM and strength measurements to ensure proper motion. The following variables were measured for all subjects in the given order: shoulder functional scale, psychological factors, and physical factors. First, the subjects were instructed to fill out the questionnaires (SPADI, DASH, VAS, EWPS, and BDI). Then, shoulder ROM (SIR, SER, SHA, and SAB) and muscle strength (SIR, SER, EF, LT, and SP) was measured in random order. The random order was determined by drawing lots. Mean values from two measurements of shoulder ROM and muscle strength were used.

### 5. Data Collection

Data from the Smart KEMA motion sensor were recorded at a sampling frequency of 25 Hz and transmitted to an Android tablet over a Bluetooth connection by using the Smart KEMA application software (KOREATECH Co., Ltd., Seoul, Korea). Moreover, data from the Smart KEMA tension sensor were recorded at a sampling frequency of 10 Hz and transmitted to an Android tablet over a Bluetooth connection by using the Smart KEMA application software. All shoulder ROM measurement data were expressed in degrees and the shoulder muscle strength measurement data were expressed in kilograms. The collected shoulder muscle strength data were normalized to body weight. Normalized strength was represented as a percentage of body weight (%BW = [maximal strength of each trial (kg)/body weight (kg)] × 100). Average strength values were calculated for subsequent analyses.

### 6. Statistical Analysis

In this study, data were expressed as mean ± standard deviation. All data were tested for normal distribution by using the Kolmogorov–Smirnov normality test. To compare the functional outcome measure scores on the SPADI, DASH, VAS, EWPS, and BDI, between-group analysis (comparison between workers with and without SIS) was performed using independent *t*-tests. Moreover, to compare shoulder ROM, muscle strength, and asymmetry ratio, the results of the between-group comparison (comparison between workers with and without SIS) were analyzed using independent *t*-tests. All analyses were carried out using SPSS 25.0 (SPSS Inc., Chicago, IL, USA). The level of significance was set to *p* = 0.05.

## Results

### 1. Functional Outcome Measure Scale

The independent *t*-tests revealed a statistically significant difference between the two groups in terms of SPADI (*p* < 0.001) and DASH (*p* < 0.01) scores, with significantly greater shoulder and upper extremity disability according to the outcome measures of the workers in the SIS group as compared to those of the workers in the without-SIS group, as can be inferred from Table 2. The workers with SIS scored significantly higher on the VAS (pain intensity) scale than the workers without SIS. However, the EWPS productivity and BDI scores of the workers in the two groups were not significantly different (*p* > 0.05).2.

**Table 1.** Subject characteristics

| **Characteristics** | **Total (n = 67)** | **Workers with**<br>**SIS (n = 35)** | **Workers without**<br>**SIS (n = 32)** |
| --- | --- | --- | --- |
| Age (years) | 45.6 ± 7.6 | 46.3 ± 7.2 | 44.8 ± 8.0 |
| Height (cm) | 171.6 ± 5.6 | 171.6 ± 5.3 | 171.5 ± 5.9 |
| Body mass (kg) | 73.2 ± 10.6 | 73.9 ± 10.4 | 72.4 ± 11.0 |

**Table 2.** Comparison of SPADI, DASH, VAS, EWPS, and BDI scores between the two groups

| | mean ± SD | | Diff. | *T* | *p* |
| --- | --- | --- | --- | --- | --- |
|  | Workers with SIS<br>(n = 35) | Workers without<br>SIS (n = 32) |  |  |  |
| SPADI | 47.78 ± 18.74 | 28.08 ± 13.95 | 19.70 | -4.845 | <0.001* |
| DASH | 32.24 ± 16.16 | 18.59 ± 17.31 | 13.64 | -3.337 | 0.001* |
| VAS (mm) | 54.43 ± 24.06 | 28.69 ± 21.74 | 25.74 | -4.579 | <0.001* |
| EWPS | 37.77 ± 10.05 | 35.59 ± 13.57 | 2.17 | -0.751 | 0.456 |
| BDI | 13.63 ± 10.21 | 11.00 ± 10.66 | 2.62 | -1.030 | 0.307 |

SPADI: Shoulder pain and disability index. DASH: Disability of arm, shoulder, and hand. EWPS: Endicott work productivity scale. BDI: Beck depression inventory.
\*Independent *t*-test *p* < 0.05.

**Table 3.** Comparison of shoulder ROM between the two groups

| | mean ± SD (°) | | Diff. | *T* | *p* |
| --- | --- | --- | --- | --- | --- |
|  | Workers with SIS<br>(n = 35) | Workers without<br>SIS (n = 32) |  |  |  |
| SIR | 43.25 ± 9.31 | 57.10 ± 15.36 | 13.84 | 4.504 | <0.001* |
| SER | 97.01 ± 8.87 | 98.88 ± 8.30 | 1.86 | 0.887 | 0.378 |
| SAB | 146.38 ± 24.84 | 162.58 ± 14.60 | 16.20 | 3.216 | 0.002* |
| SHA | 29.41 ± 9.73 | 31.21 ± 9.65 | 1.79 | 0.757 | 0.452 |

SIR: Shoulder internal rotation. SER: Shoulder external rotation. SAB: Shoulder abduction. SHA: Shoulder horizontal adduction. \*Independent *t*-test *p* < 0.05.

**Table 4.** Comparison of shoulder muscle strength between the two groups

| | mean ± SD (%BW) | | Diff. | *T* | *p* |
| --- | --- | --- | --- | --- | --- |
|  | Workers with SIS<br>(n = 35) | Workers without<br>SIS (n = 32) |  |  |  |
| SIR | 15.55 ± 5.38 | 19.17 ± 6.08 | 3.61 | 2.582 | 0.012* |
| SER | 11.20 ± 3.85 | 12.87 ± 3.46 | 1.66 | 1.861 | 0.067 |
| EF | 23.32 ± 6.55 | 25.09 ± 7.74 | 1.76 | 1.012 | 0.315 |
| LT | 7.98 ± 3.62 | 9.34 ± 3.44 | 1.35 | 1.571 | 0.121 |
| SP | 23.82 ± 7.89 | 25.54 ± 7.83 | 1.71 | 0.893 | 0.375 |

SIR: Shoulder internal rotation. SER: Shoulder external rotation. EF: Elbow flexion.
LT: Lower trapezius. SP: Shoulder protraction. \*Independent *t*-test $p < 0.05$ .

**Table 5.** Comparison of asymmetry ratios of shoulder muscle strengths between the two groups

| | mean ± SD (%AR) | | Diff. | *T* | *p* |
| --- | --- | --- | --- | --- | --- |
|  | Workers with SIS<br>(n = 35) | Workers without<br>SIS (n = 32) |  |  |  |
| SIR | 10.17 ± 17.05 | -0.62 ± 12.75 | 10.78 | -2.911 | 0.015* |
| SER | 9.05 ± 15.48 | 0.93 ± 10.38 | 8.11 | -2.497 | 0.005* |
| EF | 6.8 ± 8.93 | 1.36 ± 7.00 | 5.44 | -2.759 | 0.008* |
| LT | 4.86 ± 16.57 | 1.38 ± 15.36 | 3.47 | -0.889 | 0.377 |
| SP | 3.38 ± 10.09 | 1.16 ± 8.29 | 2.21 | -0.976 | 0.333 |

SIR: Shoulder internal rotation. SER: Shoulder external rotation. EF: Elbow flexion.
LT: Lower trapezius. SP: Shoulder protraction. \*Independent *t*-test $p < 0.05$ .

### 2. Shoulder ROM

The results of the independent *t*-test indicated significant differences in SIR and SAB ROM between the workers with and without SIS (*p* < 0.01; *p* < 0.001). However, no significant differences were found in SER and SHA ROM between the two groups (*p* > 0.05; *p* > 0.05).

### 3. Strength of Shoulder Muscles

The result of the independent *t*-test indicated that the workers with SIS had significantly lower isometric muscle strength of SIR than the workers without SIS (*p* < 0.05). However, there was no significant difference in the isometric muscle strengths of LT and SP between the workers with and without SIS (*p* > 0.05; *p* > 0.05). Moreover, the SER and EF muscle strengths of the workers in the two groups were not significantly different (*p* > 0.05; *p* > 0.05).

### 4. Asymmetry Ratio of Shoulder Muscle Strength

The asymmetry ratios of SIR and SER muscle strength were significantly higher in the workers with SIS compared to those in the workers without SIS (*p* < 0.01; *p* < 0.05). Moreover, the asymmetry ratio of EF muscle strength was significantly higher in the workers with SIS compared to that in the workers without SIS (*p* < 0.01). However, there were no statistically significant differences in the asymmetry ratios of LT and SP muscle strengths between the two groups (*p* > 0.05; *p* > 0.05).

## Discussion

According to the results, VAS was significantly higher in workers with SIS than in workers without SIS. The average SPADI score for the workers with SIS was 47.78 points, which was approximately 20 points higher than the average score for the workers without SIS. The average DASH score for the workers with SIS was 32.24 points, which was approximately 14 points higher than the average for the workers without SIS. These results indicate that the workers with SIS found it more difficult to perform upper limb movements in their daily lives than the workers without SIS. However, the EWPS (productivity) and BDI (depression severity) scores of the workers in the two groups did not differ significantly. EWPS is a productivity questionnaire that tests physical, psychological, and sociological factors, such as attendance, quality of work, performance capacity, as well personal factors, such as social, mental, physical, and emotional states. Thus, we expected that there would have be no difference between the workers with and without SIS.

Researchers have found that patients with SIS exhibit decreased SAB or flexion ROM [7,33,34]. According to Endo et al. (2001), SIS caused a significant reduction in SAB ROM. Our study also showed that the average SAB in the frontal plane for workers with SIS was 148°, which is significantly lower than the average SAB angle of 170° for workers without SIS. Joung et al. (2019) found that SIS patients exhibited decreased SHA ROM owing to shortening of the posterior deltoid muscle and tightness of posterior capsule. Also, Tyler et al. (2000) reported that patients with SIS in their dominant arm or non-dominant arm exhibited posterior capsule tightness and reduced SIR ROM compared to normal subjects. They stated that SIS patients lose SIR ROM in their involved arm because of posterior capsule tightness. Similarly, in this study, workers with SIS exhibited significantly reduced SIR ROM compared to workers without SIS. However, the present study did not find a significant difference in SHA ROM between the workers with and without SIS. Tyler et al. (2000) fixed the scapula of the arm to be measured directly with the examiner’s hand and then measured SHA ROM to examine posterior capsule tightness. However, since we measured SHA ROM with the arm to be measured on the table, we expect no difference between the workers with and without SIS. Furthermore, they reported that the SER ROM of the dominant arm of SIS patients did not decrease, but the SER ROM of the non-dominant arm of SIS patients decreased significantly. They ascribed this decrease to reduced demand on the non-dominant arm to perform day-to-day activities. Our study also showed no significant difference between the workers with and without SIS because most of the workers in the SIS group had impingement in their arms on the dominant side.

SIR muscle weakness is considered common clinical feature in SIS [7,33]. Leroux et al. (1994) found that the SIR strength of the involved arm of SIS patients in the sitting position with 45° abduction and 30° flexion was reduced by 48–53% compared to the dominant arm of the subjects in the control group. In addition, several studies have suggested that SER muscle weakness may contribute to SIS [34,37]. However, according to Erol et al. (2008), there was no difference in SER muscle strength between subjects with and without SIS. Also, according to Leroux et al. (1994), the degree of decrease in the SIR strength of SIS patients was relatively higher than the degree of decrease in SER strength. The results of our study indicate average decreases of 19% and 13% in the SIR and SER strengths of the workers with SIS, respectively, compared to those of workers without SIS. However, in this study, there was a significant difference in SIR strength between the two groups, but not a significant difference in SER strength. Cools et al. (2005) found that overhead athletes with SIS had a significant difference in the strength of SP muscle on the injured side compared with the dominant side of a healthy group. Also, Cools et al. (2007) found significantly lower LT muscle activity when overhead athletes with SIS performed isokinetic abduction at 120°/s. Unlike athletes, who mainly perform overhead activities such as throwing or wielding a tennis racket, workers often place their arms under the shoulder level and perform motions for assembly. Therefore, in this study, there would be no difference in the strengths of the SP and LT muscle, which play the major roles in scapular posterior tilt and upward rotation when raising arms, between the workers with and without SIS. Also, muscle strength varies widely based on physical characteristics, including body weight, height, shape of bone or joint, and psychological characteristics, including cognition, emotion, and motivation [40]. Moreover, shoulder muscle strength assessment performed using a maximal isometric contraction showed that the rotator cuff muscles and relatively large muscles (pectorals, deltoid, upper trapezius, rhomboid) worked together; there were no significant differences in the strengths of the scapulothoracic and glenohumeral joint muscles between the workers with and without SIS.

The main findings of this study are that the asymmetry ratios of EF, SER, and SIR are greater in the workers with SIS than those in the workers without SIS. This means that the involved arm of the workers with SIS had upper extremity muscle strength deficits for EF, SER, and SIR motions compared to the uninvolved arm. These results can be explained as follows. First, Richards et al. (2005) found that coracohumeral distance, one of the factors contributing to the development of SIS, was significantly reduced in patients requiring subscapularis repair surgery. In other words, a narrowed coracoid distance is one of the possible causes of subscapularis injury. Reddy et al. (2000) reported that subscapularis muscle activity was significantly decreased in 30°-60° of scaption in patients with SIS compared to those in the normal group. Therefore, decreased subscapularis muscle activity along with the possibility of a damaged subscapularis due to narrowed coracohumeral distance probably contributed to the high asymmetry ratio of SIR in the workers with SIS. Second, infraspinatus and teres minor fatigue among SER muscles can cause SIS by reducing the scapular posterior tilt movement during arm raising [43]. In addition, infraspinatus muscle activity was shown to be decreased between 30 and 90 degrees during scaption [42]. Although SER strength did not differ between the workers with and without SIS due to the relatively large deviations between individuals, owing to fatigue or poor infraspinatus and teres minor muscle activity, the asymmetry ratio of SER muscle strength would be significantly higher in the workers with SIS than that in the workers without SIS. In addition, we found no difference in EF muscle strength between the workers with and without SIS, but the workers with SIS exhibited significantly higher asymmetry ratios of EF muscle strength than the workers without SIS. These results can be explained as follows. First, the long head of the biceps brachii is a depressor of the humeral head that prevents SIS when it functions properly during arm elevation [4]. When this muscle lacks strength or does not function properly, the humeral head is not depressed, which increases the likelihood of SIS development. Therefore, the long head of biceps brachii, which depresses the humeral head workers, on the involved side of the workers with SIS would have lacked strength compared to that on the uninvolved side. Second, tendinitis of the long head of the biceps brachii is a common problem associated with SIS or rotator cuff tearing [44,45]. Especially, Neer (1983) reported that most biceps tendinitis is caused by SIS. Thus, pain due to biceps tendinitis associated with SIS can be thought to contribute to muscle strength deficit on the involved side of the elbow flexor compared to the uninvolved side.

There were some limitations to this study. First, since this study only covers male workers, the results of this study cannot be generalized to all genders. Second, because it is aimed at adults who are currently engaged in vocational activities, it cannot be generalized to adolescents or the elderly. Third, because the study involved workers working in assembly lines, it is not possible to generalize these results to office workers who do not have a heavy load on their shoulder or to athletes who repeatedly perform certain motions.

## Summary and Conclusion

The results of this study showed that the SPADI and DASH scores, which indicate shoulder function, were higher, and the intensity of self-aware pain was higher in workers with SIS. Also, workers with SIS had lower SIR and smaller SAB ROM. In addition, workers with SIS had lower SIR muscle strength than workers without SIS. Particularly, the asymmetry ratios of SIR, SER, EF muscle strength are good comparable factors for workers with and without SIS. The asymmetry ratios of shoulder muscle strengths could provide an important baseline comparison for the workers with SIS.

## Data Availability

Data will be made available on request.

---

**Figure 1.**
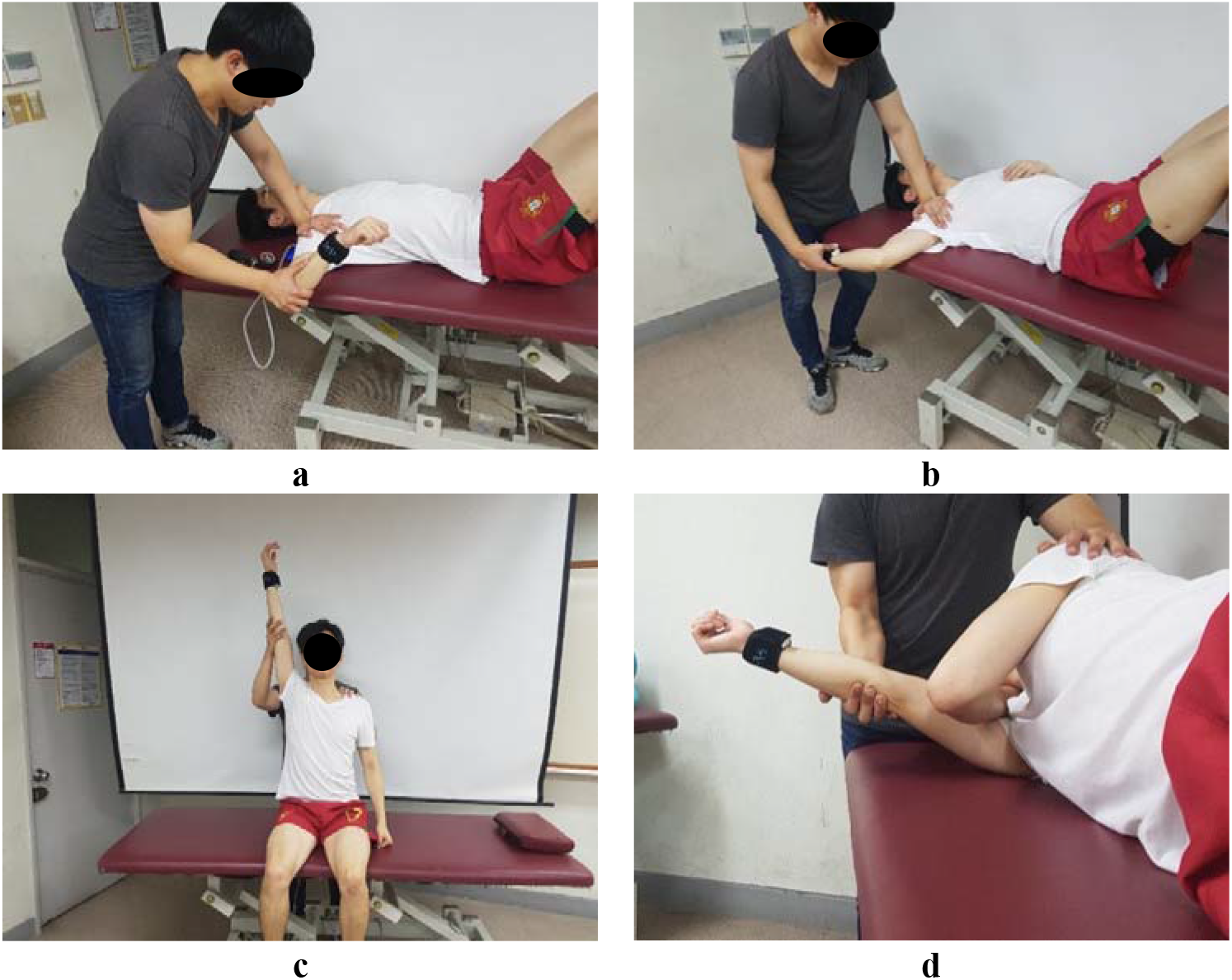
Measurement of shoulder ROM: a) shoulder internal rotation; b) shoulder external rotation; c) shoulder abduction; d) shoulder horizontal adduction

**Figure 2.**
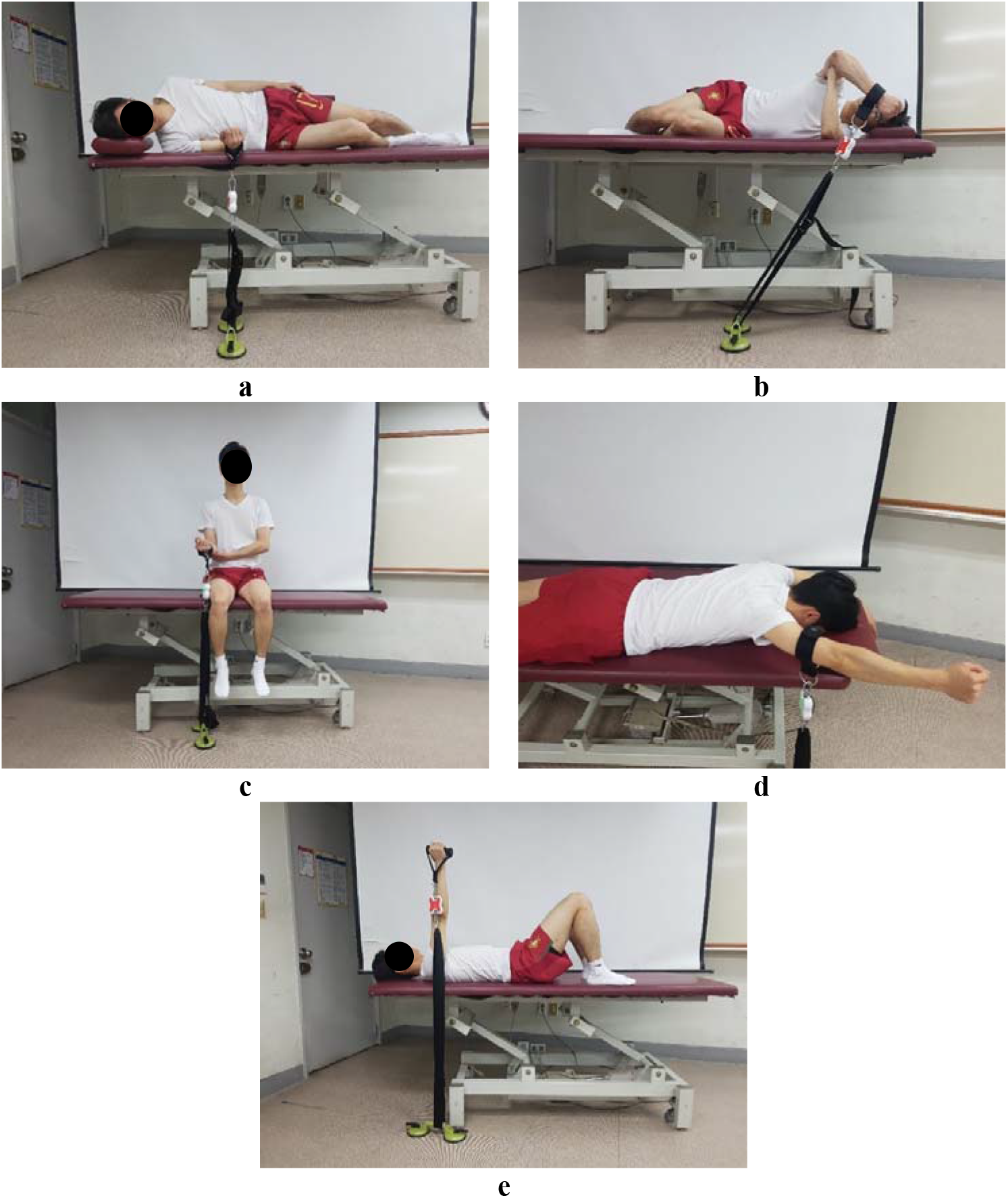
Measurement of shoulder muscle strength: a) shoulder internal rotation; b) shoulder external rotation; c) elbow flexion; d) lower trapezius; e) scapular protraction

